# Rapid evidence summary on SARS-CoV-2 survivorship and disinfection, and a reusable PPE protocol using a double-hit process

**DOI:** 10.1101/2020.04.02.20051409

**Authors:** José G B Derraik, William A Anderson, Elisabeth A Connelly, Yvonne C Anderson

## Abstract

In the COVID-19 pandemic caused by SARS-CoV-2, hospitals are stretched beyond capacity. There are widespread reports of dwindling supplies of personal protective equipment (PPE), which are paramount to protect frontline medical/nursing staff and to minimize further spread of the virus. We carried out a rapid review to summarize the existing evidence on SARS-CoV-2 survivorship and methods to disinfect PPE gear, particularly N95 filtering facepiece respirators (FFR). In the absence of data on SARS-CoV-2, we focused on the sister virus SARS-CoV-1. We propose a two-step disinfection process, which is conservative in the absence of robust evidence on SARS-CoV-2. This disinfection protocol is based on an initial storage of PPE for ≥4 days, followed by ultraviolet light (UVC), dry heat treatment, or chemical disinfection. Importantly, each of the two steps is based on independent disinfection mechanisms, so that our proposed protocol is a multiplicative system, maximising the efficacy of our disinfection process. This method could be rapidly implemented in other healthcare settings, while testing of each method is undertaken, increasing the frontline supply of PPE, and avoiding many of the upstream issues of supply chain disruption currently being faced.

## 1. INTRODUCTION

In pandemic situations, such as the current COVID-19 scenario, hospital resources are frequently stretched beyond capacity, as has already occurred in many countries across the globe^1^. Preventing the spread of COVID-19 to and from health care workers and patients relies on the availability and effective use of personal protective equipment (PPE)^1^. PPE includes masks, eye protection, gloves, gowns, and in the event of aerosol-generating procedures, N95 filtering facepiece respirators (FFR) or FFP2 standard (or equivalent)^2^.

Two small studies from the same group in Singapore failed to detect SARS-CoV-2 contamination from PPE, but only on the surface of uncovered shoes^3,4^. In the case of SARS-CoV-2, it has been estimated that 3.2% of patients in China required intubation^5^. Evidence from the SARS-CoV-1 epidemic show that doctors and nurses involved in the early critical care period and endotracheal intubation of patients were more than 13 times more likely to acquire SARS-CoV-1 infection themselves^6^. Given this results in a significant loss of highly specialised healthcare workers in an already strained workforce, avoidance of cross contamination is critical in all health care settings.

The World Health Organization acknowledges the current global stockpile is insufficient, particularly for masks and respirators^2^, and supply of gowns and goggles is also expected to be insufficient. Coordinating the supply chain of PPE in the midst of an epidemic with many closed borders and reduced freight is challenging. Individual behaviour becomes a factor when people are scared or ill-informed^7^, theft of PPE can occur, and local supply chain issues mean that inappropriate use of PPE happens due to lack of supply, despite best-practice guidance on its use^2^. A call for ideas on conserving PPE was made through JAMA on 20 March 2020^8^. One recommendation was reusing PPE. Given the ability of this to rapidly increase supply issues close to the frontline avoiding many of the upstream disruption to the supply chain, this rapid evidence summary was prepared, aiming in particular at the re-utilization of the usually disposable N95 FFRs during the current epidemic. A whole-of-PPE solution has been developed within this protocol, which would be able to be rapidly set up in many healthcare settings.

## 2. VIRUS SURVIVORSHIP

In light of the very recent identification of SARS-CoV-2, there is a near complete lack of data on the survival of this virus in the environment under different conditions, as well as efficacy of disinfection methods. In the absence of data on SARS-CoV-2, we have focused on SARS-CoV-1, which forms a sister clade virus from the same species^9^.

### 2.1. SARS-CoV-1

- Materials tested included cardboard^10^, wood^11^, plastic^11-14^, fabric^11,12^, paper^11,12^, and metal^10,11^ (Table 1).
- Survival on a range of materials varied somewhat, and even within type (e.g. stainless steel vs copper^10^).
- One study demonstrated survival of 2 days on a disposable polypropylene gown and 24 hours on a cotton gown^12^ (Table 1).
- Survival at room temperature and at 40 to 50% relative humidity was as long as 9 days on a polystyrene petri dish^13^ and approximately 21 days also on plastic^14^ (Table 1).
- 9-day survival in respiratory specimens at room temperature^12^, but more than 14 days in dechlorinated tap water at 4°C and >17 days in urine at 20°C^15^ (Table 1).
- Of note, a study with a surrogate coronavirus (i.e. transmissible gastroenteritis virus) showed that this virus was detectable on N95 respirators for up to 24 hours^16^.
- It is important to highlight the effect of inoculum size on SARS-CoV-1 inactivation, as clearly shown by Lai et al. 2005^12^. While inoculation of a cotton gown at 10^4^ TCID_50_/ml led to inactivation in 5 minutes, at 10^6^ TCID_50_/ml inactivation took 24 hours.

### 2.2. SARS-CoV-2

- Only one study found^10^. Virus undetectable after 4 days on plastic and stainless steel, with shorter survival on cardboard and copper^10^ (2 days and 4 hours, respectively) (Table 1).

### 2.3. Survivorship summary

- It would be ill-advised to rely on a single study on SARS-CoV-2 to draw any clear conclusions on the virus’ survival on different surfaces, especially in light of the existing data on SARS-CoV-1. Thus, until new evidence comes to light, it should be assumed that SARS-CoV-2 may survive for much longer periods under certain conditions, as shown to be the case for SARS-CoV-1.
- Unless PPE needs to be immediately re-used, PPE that is visually intact and visually clean could be stored for a stand-down period of **at least** 4 days before undergoing a given disinfection treatment (i.e. ensuring the virus receives a double-hit).

## 3. DISINFECTION

A wide variety of disinfection methods for PPE have been examined and reported in the literature. These can be characterized as either 1) energetic methods (e.g. ultraviolet, dry and wet heat, and microwave generated steam), or 2) chemical methods (e.g. alcohol, ethylene oxide, bleach, and vapourized hydrogen peroxide). Some of these rapidly and significantly affect N95 filter performance (alcohol^17^), and others require chemical supplies and specialized facilities (e.g. ethylene oxide, vapourized hydrogen peroxide), or are not readily scalable to large numbers of PPE (e.g. microwave generated steam). We focus here on methods that may be easier to implement at a useful scale.

**TABLE 1.** Studies reporting on the survival of SARS-CoV-1 and SARS-CoV-2.

| Study | Virus | Inoculum & conditions | Material & result |
| --- | --- | --- | --- |
| Duan 2003 <sup>11</sup> | SARS-CoV-1<br>[strain P9] | 10<sup>6</sup> TCID<sub>50</sub> in 300 µl<br>Room temperature (~20°C) | Time to undetectable CPE:<br>Wood board, Mosaic 4 days<br>Glass, press paper, plastic, water, soil 5 days<br>Metal, cloth, filter paper – few cells still detected after 5 days<br>Serum, filtrated sputum 4 days<br>Sputum, faeces, filtrated faeces, urine – cells still detected after 5 days |
| Lai 2005 <sup>12</sup> | SARS-CoV-1 | 10<sup>6</sup> TCID<sub>50</sub>/ml | Time to inactivation:<br>Disposable polypropylene gown 2 days<br>Cotton gown 24 hours<br>Paper 24 hours<br>Respiratory specimens at room temperature 9 days |
| Rabenau 2005 <sup>13</sup> | SARS-CoV-1 | 500 µl virus suspension applied to dish and left to dry at 21-25°C, unknown RH | Plastic (polystyrene petri dish) – infectivity only lost after 9 days.<br>In suspension remained infective after 9 days. |
| Wang 2005<sup>15</sup> | SARS-CoV-1 | 1 ml of 10<sup>5</sup> TCID<sub>50</sub>/ml | •Time to no detection at 20°C:<br>Hospital wastewater, domestic sewage, dechlorinated tap water 3 days<br>PBS >14 days<br>Stool 4 days<br>Urine >17 days<br>•Time to no detection at 4°C:<br>Hospital wastewater, domestic sewage, dechlorinated tap water, PBS >14 days |
| Chan 2011 <sup>14</sup> | SARS-CoV-1 | 10<sup>5</sup> TCID<sub>50</sub>/ 10 µl<br>22-25°C, 40-50% RH | Plastic – lost viability after ~21 days |
| van Doremalen 2020 <sup>10</sup> | SARS-CoV-1 | 10<sup>5</sup> TCID<sub>50</sub>/ml<br>21-23°C, 40% RH<br>Surface deposits of 50 µl | Time to limit of detection*<br>Plastic 4 days<br>Stainless steel 3 days<br>Cardboard 24 hours<br>Copper 24 hours |
| van Doremalen 2020 <sup>10</sup> | SARS-CoV-2 | 10<sup>5</sup> TCID<sub>50</sub>/ml<br>21-23°C, 40% RH<br>Surface deposits of 50 µl | Time to limit of detection*<br>Plastic 4 days (HL 6.8 hours)<br>Stainless steel 4 days (HL 5.6 hours)<br>Cardboard 2 days<br>Copper 8 hours |

CPE, cytopathic effect; HL, half-life; PBS, phosphate-buffered saline; RH, relative humidity; TCID<sub>50</sub>, median tissue culture infectious dose, corresponding to the concentration at which 50% of the experimental cells are infected after inoculation.
\* 10<sup>0.5</sup> TCID<sub>50</sub> per ml of medium for plastic, steel, and cardboard; 10<sup>1.5</sup> TCID<sub>50</sub> per ml of medium for copper.

### 3.1. Ultraviolet germicidal irradiation (UVGI)

Across the ultraviolet (UV) light spectrum, there are three classifications: UVA (320-400 nm), UVB (280-320 nm), and UVC (200-280 nm)^18^. UVC light has much stronger germicidal properties than both UVA and UVB^19,20^. UVC is strongly absorbed by RNA and DNA bases leading to molecular structural damage via a photodimerization process. This results in virus inactivation, such that they are no longer able to replicate^20,21^. Thus, the focus of this protocol has been on UVC.

- Studies were found only for SARS-CoV-1 (Table 2). It should be noted that those studies were almost invariably performed on aqueous solutions, in air, or on solid surfaces, i.e. environments that do not directly reflect for example, the micro-environment of N95 FFRs. As a result, the reported applied doses are at best a relative guidance.
- Ultraviolet light C (UVC) applied doses varied markedly from 300 to 14,500 mJ/cm^2^, with mixed outcomes (Table 2).
- At 360 mJ/cm^2^, SARS-CoV-1 had the highest UV D_90_ (i.e. required applied dose for 90% inactivation) among nearly 130 viruses from hundreds of published studies summarized by Kowalski^22^. In addition, in protein medium, an applied dose of 14,500 mJ/cm^2^ did not completely inactivate the virus^18^ (Table 2), due to competitive absorption of UV photons by the protein.
- UVC is effective against SARS-CoV-1, but efficacy of the applied dose (a function of irradiance and time) appears to be highly dependent on many factors, such as inoculum size, culture medium, and shape and type of material^12,20,23,24^, likely explaining the highly inconsistent findings in the published literature.
- Based on the available evidence it seems that the effect of relative humidity on UVGI efficacy can be considered negligible^22^.
- Importantly, the applied dose is not necessarily the same as the actual dose the treated virus receives. While the applied dose is easy to measure experimentally, the received dose is not. If there are shadowing or absorption effects from the surrounding medium, the actual dose reaching the virus will be lower.
- The penetration of UV across the multiple layers of an N95 FFR may vary from one model and manufacturer to another^25^. There is some limited evidence that the majority (approximately 90%) of captured aerosols occurs on the outer filter layer on an N95 FFR^26^. Therefore, providing a larger UV dose on the outside surface may be desirable.
- Overall, the effective applied dose is unclear, but appears to be high in comparison to other viruses.
- Mills et al. (2018) reported on a more extensive set of tests on N95 masks using H1N1 viruses, and included the effects of soiling agents (artificial saliva and/or skin oil) that could reduce the efficacy of UV exposure^27^. Fifteen different N95 models were tested from a variety of manufacturers, and both the facemask and straps were monitored. All FFRs were disinfected to a level of at least 3 log (i.e. 99.9%), even in the presence of soiling agents, when the UV dose was 1,000 mJ/cm^2^. Similarly, Heimbuch & Harnish (2019) showed complete disinfection of SARS-CoV-1 from FFR coupons in the presence of artificial saliva (mucin) and artificial skin oil (sebum)^23^.
- We estimate that the minimum applied UVC dose for effective deactivation of SARS-CoV-2 on N95 FFRs would likely be close to 1,000 mJ/cm^2^, particularly in light of the mask’s porous surface (as compared to a smooth surface material), as shown by Heimbuch & Harnish’s 2019 study^23^.
- Note that the studies showing SARS-CoV-1 survival at higher doses were most likely confounded by the aqueous media (often with added protein), which would absorb UVC photons, reducing the actual dose reaching the virus.

### 3.2. Heat treatment

Heat treatment is one of the most common methods for virus deactivation. Heat induces structural changes in virus proteins, disrupting the specific structures necessary to recognize and bind to host cells^28^.

**TABLE 2.** Studies reporting on the efficacy of ultraviolet (UV) light or heat treatment against SARS-CoV-1.

| Study | Virus | Inoculum & conditions | UV exposure | UV findings & applied dose | Heat treatment | Notes |
| --- | --- | --- | --- | --- | --- | --- |
| Duan 2003 <sup>11</sup> | SARS-CoV-1 [strain P9] | 10<sup>6</sup> TCID<sub>50</sub> in 100 µl culture medium in well plates | 260 nm-length UVC<br>Irradiance: >90 µW/cm<sup>2</sup><br>Distance: 80 cm | Cell culture exposure – undetectable CPE at 60 min (*D*=300 mJ/cm<sup>2</sup>) | Undetectable CPE after:<br>30 min at 75°C<br>60 min at 67°C<br>90 min at 56°C<br>Note that RH was not reported. |  |
| Ansaldi 2004 <sup>52</sup> | SARS-CoV-1 | "standard concentration of cell-grown virus" in 1 ml salt solution on a plate<br>18°C, 40% RH | Irradiance: 40 mW/cm<sup>2</sup><br>UV type and distance to light undisclosed. | Negative result by cell culture and PCR at 5 min (*D*=12,000 mJ/cm<sup>2</sup>) | n/a | Methods poorly described. |
| Darnell 2004 <sup>19</sup> | SARS-CoV-1 | UV: 2-ml aliquots of virus in well plates<br>Heat: 320 µl in 1.5-ml polypropylene tubes cryotubes (RH undisclosed) | UVC 254 nm<br>Distance from source: 3 cm from bottom of wells<br>Irradiance: 4,016 µW/cm<sup>2</sup> | Virus inactivated to detection limit after 15 min (*D*=3,600 mJ/cm<sup>2</sup>) | Virus inactivated below limit of detection after:<br>90 min at 56°C and 65°C<br>45 min at 75°C | UVA treatment was also looked at, but found to be ineffective<br>Heat treatment reduced virus to very low levels in a shorter period of time (20 min at 56°C and 4 min at 65°C), but failed to completely inactivate it until 90 minutes. |
| Rabenau 2005 <sup>13</sup> | SARS-CoV-1 | 500 µl solutions with virus<br>unknown RH | n/a | n/a | • 56°C for 30 min reduced virus titre below detection limit, but this did not happen in presence of protein additive (20% FCS).<br>• 60°C for 30 min eliminated infectious virus, regardless of protein additive. |  |
| Darnell 2006 <sup>18</sup> | SARS-CoV-1 | UV: Virus solution in well plates<br>Heat: samples incubated in heated water bath<br>Undisclosed RH | UVC 254 nm<br>Distance from source: 3 cm from the bottom of wells<br>Irradiance: 4,016 µW/cm<sup>2</sup> | • UVC inactivated virus in PBS solution to the limit of detection by 40 min (*D*=9,600 mJ/cm<sup>2</sup>)<br>• UVC did **not** fully inactivate the virus in BSA protein solutions after 60 min (*D*=14,500 mJ/cm<sup>2</sup>) | Virus inactivated to detection limit in:<br>Human serum: 56°C for 20 min / 65°C for 10 min<br>Protein solutions: 60°C for 30 min (at highest protein content) | Study specific to non-cellular blood products |
| Kariwa 2006 <sup>31</sup> | SARS-CoV-1 | UV: 2 ml aliquots on open plastic petri dishes<br>Heat: aliquots of virus solution placed in 50-ml tubes and heated in water bath | UV "normal biosafety cabinet UV lights"<br>Distance ?<br>Irradiance: 134 µW/cm<sup>2</sup> | Failed to completely eliminate virus after 60 min (*D*=500 mJ/cm<sup>2</sup>) | No virus infectivity detected after 60 min at 56°C |  |
| Heimbuch 2019<sup>23</sup> | SARS-CoV-1 | FFR coupons in 3 soiled conditions: no soiling agent, artificial saliva (mucin) and artificial skin oil (sebum) | UVC lamp (254 nm)<br>Distance 15.2–22.9 cm<br>Irradiance: mean 2.3 mW/cm<sup>2</sup> | No detectable viable virus in the 3 conditions tested at 1,000 mJ/cm<sup>2</sup>, but UVGI was ineffective at lower applied doses. | n/a |  |

BSA, bovine serum albumin; CPE, cytopathic effect; *D*, UV applied dose; PBS, phosphate-buffered saline; RH, relative humidity; TCID<sub>50</sub>, median tissue culture infectious dose, corresponding to the concentration at which 50% of the experimental cells are infected after inoculation.
*D* was calculated by the author using the standard formula (with time expressed in seconds): $mJ/cm^2 = mW/cm^2 * time$

- Studies found only for SARS-CoV-1 (Table 2). It should be noted that it is not easy to extrapolate the results from most heat treatment studies reported here. They were often performed with the virus exposed while in solutions, which are mechanistically different from surface contamination, as one would most likely encounter on PPE that is not heavily soiled, particularly N95 FFRs.
- Environments with lower temperatures seem to be more favourable for virus survival and increased transmission rates^14,29,30^. While the efficacy of heat treatment appears to be affected by relative humidity^14^, this relationship for SARS-CoV-1 is unclear, as almost all reported experimental studies failed to report on relative humidity (Table 2). However, the association between temperature and relative humidity was not monotonic for other coronaviruses, with virus survival lowest at moderate relatively humidity (50%)^30^.
- Two studies reduced SARS-CoV-1 to below levels of detection with exposure to 56°C at 20 min^18^ and 60 min^31^, but one study showed that at 56°C for 30 min heat treatment was ineffective^13^ in protein medium (Table 2).
- Heat treatment at 60°C for 30 minutes inactivated SARS-CoV-1 in two studies^13,18^, irrespective of protein concentrations. However, in Duan et al. (2003)^11^ the virus was only inactivated at 67°C after 60 minutes, while in Darnell et al.^19^ inactivation occurred only after 90 minutes at 65°C (Table 2).
- Overall, heat treatment at 60°C for 90 minutes would reduce SARS-CoV-1 to below levels of detection according to all five heat-treatment studies reported in Table 2.

### 3.3. Disinfection summary

- The applied UVC dose should be at least 1,000 mJ/cm^2^, but we recommend an initial conservative dose of 2,000 mJ/cm^2^ [applied to each side of N95 FFRs, i.e. wearer-facing and outer sides] to account for possible errors in applied dose estimation, effects of different materials, the challenge to reach the inner filtering layers of FFRs^25^, as well as the uncertainty regarding the actual susceptibility of SARS-CoV-2 to UVGI.
- We also recommend heat treatment at 60°C for 90 minutes to treat PPE. This is most likely a very conservative protocol when applied to surface contamination, but in the absence of more precise data, we recommend erring on the side of caution. The 90-minute period of exposure is advisable to ensure there is adequate heat transfer to the inner layers of the FFRs, particularly if a number of masks are being treated at the same time (in which case we would caution against stacking them).
- While we cannot recommend a target relative humidity due to the paucity of data for SARS-CoV-1, moderate levels are likely to be more desirable (i.e. 40% to 50%).
- We advise against attempts to disinfect and reuse soiled PPE, as studies using both UVGI and heat treatment show a protective effect of protein and aqueous substrata on SARS-CoV-1 survival.
- Unpublished experimental data from our group showed that there is minimal UVC radiation on the wearer-facing side of N95 FFRs when the outer side is irradiated (outer 7.34 mW/cm^2^ vs inner 0.10 mW/cm^2^). There are reports of widespread SARS-CoV-2 infection among frontline medical staff^32^, thus, it has to be assumed that SARS-CoV-2 contamination of N95 FFRs would likely occur on both sides, particularly when there is strong evidence of asymptomatic transmission^33-35^. Therefore, we strongly recommend that both wearer-facing and outer sides of N95 masks be equally treated at the recommended UVC dose.

## 4. IMPACT OF DISINFECTION ON N95 FFRs

### 4.1. UVGI

- Table 3 summarizes six studies that have looked at the effects of UVGI disinfection on the performance and structure of N95 FFRs.
- In five studies, applied doses varied somewhat from 180 mJ/cm^2^ to 6,900 mJ/cm^2^, but there were no observed effects on the N95 FFRs filter aerosol penetration, filter airflow resistance, fit, odour detection, comfort, donning difficulty, or physical appearance (Table 3).
- Heimbuch & Harnish 2019^23^ evaluated the effects of multiple UVGI cycles on 15 different N95 FFR models. Up to 20 UVGI cycles (total applied UVC dose 20,000 mJ/cm^2^) did not have a meaningful effect on fit, airflow resistance, or particle penetration for any model. Strap strength was unaffected by 10 UVGI cycles (total applied dose 10,000 mJ/cm^2^), but 20 cycles had some effect on certain models.
- Lindsley et al.^36^ went further, estimating the cumulative effect of extremely high exposures of N95 FFRs to UVC in order to mimic repeated cycles of UVGI treatment. Their lowest applied dose of 120,000 mJ/cm^2^ reduced the bursting strength of the four N95 models tested by 11% to 42% (depending on the model and the individual layer), with very minor effects on filter aerosol penetration and filter airflow resistance (Table 3). An applied dose of 590,000 mJ/cm^2^ reduced the breaking strength of straps from the four N95 FFR models tested by 10% to 21%^36^. It should be noted that their lowest dose is 120,000 mJ/cm^2^, which is 60 times higher than the conservative minimum dose of 2,000 mJ/cm^2^ we recommend for SARS-CoV-2 inactivation.

### 4.2. Heat treatment

- Table 4 describes five studies that examined the effects of heat treatment on the performance and structure of N95 FFRs.
- Two studies looked at dry heat treatment at 80°C^17,37^, reporting no meaningful effects on filter particle penetration and leading to no obvious signs of damage (Table 4).
- Three studies looked at moist heat incubation at 60°C and 80% relative humidity, two for 15 minutes^38,39^ and one for 30 minutes^40^ (Table 4) – there were no meaningful effects on filter aerosol penetration or filter airflow; most FFRs were undamaged, but in the three studies there was separation of the inner foam nose cushion from the FFR body in one particular model.
- While two studies have looked at the effects of 3 heat treatment cycles for 15 minutes^38^ and 30 minutes^40^ at 60°C on N95 FFRs, no peer-reviewed studies seem to have looked at the potential effects of more than 3 heat treatment cycles or multiple cycles of longer duration on N95 FFRs. The exception is Liao et al. (2020)^41^, who have recently reported that 20 cycles of dry heat at 75°C for 30 minutes did not affect the filtration efficacy of the key fabric in N95 FFRs. However, due to their methodology it was not possible to ascertain whether the fit of the masks for example, would be affected by their treatment protocol.
- It is worth noting that N95 FFRs are mostly made of polypropylene^42^, whose maximum operating temperature is approximately 80°C^43^, so that heat treatment approaching this temperature is probably ill-advised.

### 4.3. Summary on the impact of disinfection on N95 FFRs

- We recommend the use of UVGI at a conservative applied UVC dose 2,000 mJ/cm^2^ (for each surface) for N95 FFRs.
- Based on the available evidence, there are uncertainties about using our recommended heat treatment at 60°C for 90 minutes for N95 FFRs, as the extended time required for heat treatment may have adverse effects that could compromise its safety for re-use, especially after multiple disinfection cycles. However, a recent report by Liao et al.^41^ suggests that our proposed heat treatment regimen could be applied to N95 FFRs, and could therefore be adopted in the absence of UVC treatment.

## 5. DISINFECTION OF OTHER PPE

- Apart from N95 FFRs, in a pandemic situation the supply of other PPE will be seriously affected, including isolation gowns, surgical masks, face shields, and goggles.
- Heat treatment (at 60°C for 90 minutes) is recommended for isolation gowns (due to their size and folds) and surgical masks (due to their folded construction).
- Face shields are made of thin plastic, and usually have a foam-like material on the area that is in direct contact with the face, which would be difficult to clean with chemical disinfectants. As the shields may be damaged at 60°C we therefore recommend that these are treated with UVGI. However, repeated UVGI treatment could affect the clarity of the shields leading to ‘fogging’, in which case they should be discarded.
- Goggles and other eyewear should be immersed for at least 10 minutes in a chlorine solution at a conservative dose of 5,000 mg/l, which would account for the gradual reduction in chlorine concentration throughout the day. Alternatively, these could be cleaned with an alcohol solution at ≥80%, which should be left for at least 30 seconds^44^. Afterwards, the goggles/eyewear should be rinsed thoroughly with warm water to remove the disinfectant solution, which could otherwise damage the equipment or cause skin irritation on the wearer. Also, as goggles and other eyewear can be made of different materials, we recommend testing to make sure the disinfectant would not damage them (e.g. ‘fogging’ the lenses) before implementing a chemical disinfection procedure.

## 6. CAUTIONARY NOTES

### 6.1. Reuse of N95 FFRs

- Re-use of FFRs is not encouraged if at all possible, as high levels of disinfection cannot be guaranteed for all FFRs under all circumstances.
- According to the US Centers for Disease Control and Protection (CDC), it is not possible to determine a maximum possible generic number of safe re-uses for N95 FFRs^45^.
- CDC recommend that in the absence of manufacturer’s guidance, N95 FFRs should not be re-used more than 5 times^45^, as suggested by Fisher & Shaffer 2014^42^ and Bergman et al. 2012^46^ based on the subsequent reduction of FFRs fit.

### 6.2. Extended use of N95 FFRs

- According Fisher & Shaffer 2014^42^ extended use is preferable over limited re-use due to a lower risk of contamination with lesser contact with FFR surface.
- However, extended used leads to an increase in non-compliant behaviours (e.g. adjusting or touching the N95) over time^47^, increasing the risk of self-contamination.
- 97% of 542 first-line healthcare workers in China during the COVID-19 response had some form of skin damage, which increased with longer wear of N95 FFRs^48^. An accompanying editorial highlighted that this increases the likelihood of non-compliant FFR-wearing behaviour, and consequently an increased risk of viral transmission^49^.
- As prolonged skin breakdown increases health care workers susceptibility to infection and improper PPE use, access to virtual dermatology clinics for healthcare workers is strongly recommended to manage and treat skin breakdown in health professionals wearing PPE for extended periods.

### 6.3. Alcohol

- Due to the widespread use of alcohol-base disinfectants, it is important to emphasise that masks and respirators should not be sprayed with alcohol. Alcohol can remove the electrostatic charge from the respirator filter material, severely reducing the filter’s effectiveness at collecting particles, as shown by a number of studies^17^.

## 7. RECOMMENDATIONS

Given the dearth of evidence of PPE disinfection in SARS-CoV-2, our recommendations have been conservative, and have concluded a double-hit process would be favourable to one, until robust evidence on the efficacy of individual methods against SARS-CoV-2 is available. Importantly, each of the two steps is based on two different and independent disinfection mechanisms. Therefore, our dual-step disinfection protocol is a multiplicative process, where if each step can achieve a 3-log reduction, the two consecutive steps would theoretically achieve a 6-log reduction in SARS-CoV-2 (i.e. 99.9999%).

**TABLE 3.** Studies reporting on the effects of ultraviolet germicidal irradiation (UVGI) on N95 filtering facepiece respirators.

| Study | Treatment details | N95 | Key findings |
| --- | --- | --- | --- |
| Viscusi 2007 <sup>17</sup> | Laminar flow cabinet with a 40 W UVC light (254 nm)<br>Irradiance of 0.24 mW/cm<sup>2</sup><br>Treatment 1: 30 min, total applied dose 400 mJ/cm<sup>2</sup> [200 mJ/cm<sup>2</sup> per side (i.e. inner & outer)]<br>Treatment 2: 8 hr, total applied dose 6,900 mJ/cm<sup>2</sup> (3,450 mJ/cm<sup>2</sup> per side) | 1 unidentified N95 FFR model | Average filter particle penetration not significantly affected by either treatment.<br>No "significant visible changes" observed for any samples after either treatment. |
| Viscusi 2009 <sup>37</sup> | Laminar flow cabinet with a 40 W UVC light (254 nm)<br>Average irradiance 0.18 to 0.20 mW/cm<sup>2</sup><br>15 min exposure to each side (outer and inner)<br>Total applied dose ~180 mJ/cm<sup>2</sup> per side | Not identified by the authors, but included 3 N95 FFRs and 3 surgical N95 respirators | No effect on filter aerosol penetration, filter airflow resistance, or physical appearance |
| Bergman 2010 <sup>40</sup> | UVC lamp 40 W (254 nm)<br>45-min exposure at 1.8 mW/cm<sup>2</sup> (total applied dose 4,900 mJ/cm<sup>2</sup>)<br>Distance ~25 cm<br>Only the exteriors of the FFRs were exposed | Authors reported using the same identified equipment as those in Viscusi 2009<sup>37</sup>, i.e. 3 N95 FFRs and 3 surgical N95 respirators | UVGI-treated samples had expected levels of filter aerosol penetration (<5%) and filter airflow resistance.<br>UVGI-treated samples had similar mean % penetration to the treated samples tested in Viscusi 2009<sup>37</sup> at much lower applied doses<br>There were no observed physical damage to the FFRs |
| Bergman 2011 <sup>38</sup> | Laminar flow cabinet with a 40 W UVC lamp (254 nm)<br>Irradiance of 1.8 mW/cm<sup>2</sup><br>15 min exposure to outer FFR side (total applied dose 1,600 mJ/cm<sup>2</sup>) | 3 models tested:<br>3M 1860, 3M 1870, and Kimberly Clark PFR95-270 | There were no significant changes in FFR fit.<br>There were no observed physical damage to the FFRs |
| Viscusi 2011 <sup>39</sup> | Laminar flow cabinet with a 40 W UVC lamp (254 nm)<br>Irradiance of 1.8 mW/cm<sup>2</sup><br>Total exposure 30 min (15 min inner side and 15 min outer side)<br>Applied dose 1,600 mJ/cm<sup>2</sup> per side | 6 models: 3M 8000, 3M 8210, Moldex 2200, 3M 1860, 3M 1870, and Kimberly Clark PFR95-270 | Authors concluded that UVGI unlikely to lead to significant changes in fit, odor detection, comfort, or donning difficulty. |
| Lore 2012 <sup>53</sup> | Laminar flow cabinet, with dual-bulb 15-W UVC lamp (254 nm), 25 cm above surface<br>Irradiance 1.6 to 2.2 mW/cm<sup>2</sup><br>Total exposure 18 kJ/m<sup>2</sup> (i.e. 1,800 mJ/cm<sup>2</sup>) over 15 min | 3M 1860s, 3M 1870 | There was no significant decrease in filter performance |
| Lindsley 2015 <sup>36</sup> | UVC (254 nm)<br>91 x 31 x 64 cm chamber<br>~27°C at 25% relative humidity<br>Test pieces:<br>Respirator coupons: 0, 120, 240, 470, 710, or 950 J/cm<sup>2</sup> of UVC on each side (one side was exposed at a time)<br>Respirator straps: 0, 590, 1180, or 2360 J/cm<sup>2</sup> | 4 models tested:<br>3M 1860, 3M 9210, Gerson 1730, and Kimberly-Clark 46727 | Slight decrease in particle penetration, estimated as up to ~1 percentage point.<br>Small increase in flow resistance (<6% of the original value), independent of applied UV dose.<br>At ≥710 J/cm<sup>2</sup> there was major loss of bursting strength for most respirator layers tested, some as much as 90%. For some layers of models 3M 9210 & K-C 46727 loss >80% occurred at 470 J/cm<sup>2</sup>.<br>At 590 J/cm<sup>2</sup> the mean strap breaking strengths decreased by 10–21%<br>The lowest applied dose tested of 120 J/cm<sup>2</sup> reduced the bursting strength of the four models tests by 11% to 42% (depending on layer and model). |
| **Heimbuch 2019** <sup>23</sup> | UVC (254 nm)<br>10 or 20 cycles of 1,000 mJ/cm<sup>2</sup>, i.e. total applied doses of 10.0 or 20.0 J/cm<sup>2</sup> per FFR, respectively | 15 models tested* | Up to 20 cycles of UVGI treatment (20 J/cm<sup>2</sup>) did not have a meaningful effect on fit, air flow resistance, or particle penetration for any model. Strap strength was unaffected data by 10 UVGI cycles, but 20 cycles had some effect on certain models. |
| **Liao 2020** <sup>41</sup> | Sterilizer cabinet 8-W bulb UVC (254 nm)<br>Irradiance not described<br>10 cycles of 30 minutes | 15 x 15 cm pieces of meltblown fabric, described as the most important layer of N95 FFRs<sup>41</sup> | • This is an unpublished report that has not been peer-reviewed, with poorly described methods.<br>• The ten 30-minute cycles did not affect the fabric's filtration efficiency<br>• However, in the absence of any information on the irradiance, it is not possible to ascertain the actual applied UVC dose. |

FFR, filtering facepiece respirators, UVC, ultraviolet light C.
\* 10 cycles: 3M 1860, 3M 1870, 3M VFlex 1805, Alpha Protech 695, Gerson 1730, Kimberly-Clark PFR, Moldex 1512, Moldex 1712, Moldex EZ-22, Precept 65-3395, Prestige Ameritech RP88020, Sperian HC-NB095, Sperian HC-NB295, US Safety AD2N95A, and US Safety AD4N95; 20 cycles: 3M 1860, 3M 1870, 3M VFlex 1805, Kimberly-Clark PFR, Moldex 1512, and US Safety AD4N95.

**TABLE 4.** Studies reporting on the effects of heat treatment on N95 filtering facepiece respirators.

| Study | Treatment details | N95 | Key findings |
| --- | --- | --- | --- |
| Viscusi 2007<sup>17</sup> | Dry heat in laboratory oven<br>Treatment 1: 80°C for 60 min, with mask turned over at 30 min<br>Treatment 2: 160°C for 60 min, with mask turned over at 30 min | 1 unidentified N95 FFR model | • At 80°C, there was a small increase (negligible) in average filter particle penetration<br>• At 80°C, there were no visible changes after 60 minutes<br>• At 160° C, FFRs largely melted |
| Viscusi 2009<sup>37</sup> | Dry heat in laboratory oven<br>Treatment for 1 hour at 80°C, 90°C, 100°C, 110°C, and 120°C | Not identified by the authors, but included 3 N95 FFRs and 3 surgical N95 respirators | • Results are difficult to interpret, but it seems that the models tested maintained their expected aerosol filtration efficiency at 80°C and 90°C, without any evident signs of damage. |
| Bergman 2010<sup>40</sup> | 3 cycles of moist heat incubation<br>30-min incubation at 60°C, 80% RH in laboratory incubator<br>After 1<sup>st</sup> incubation, samples were removed from incubator and air-dried overnight. After 2<sup>nd</sup> and 3<sup>rd</sup> incubations, samples were removed from incubator and air-dried for 30 min using a fan | Not identified by the authors, but included 3 N95 FFRs and 3 surgical N95 respirators | • Heat-treated samples had expected levels of filter aerosol penetration (<5%) and filter airflow<br>• Treatment caused all samples of one FFR model to experience partial separation of the inner foam nose cushion from the FFR. |
| Bergman 2011<sup>38</sup> | Moist heat incubation (MHI)<br>15 min at 60°C (upper temp. limit), 80% RH | 3M 1860, 3M 1870, and Kimberly Clark PFR95-270 | • There were no significant changes in FFR fit.<br>• 3M 1870 samples experienced a slight separation of the inner foam nose cushion (some to a lesser or greater degree) from the FFR body, but multiple treatments did not appear to increase the level of separation compared to a single treatment. |
| Viscusi 2011<sup>39</sup> | Moist heat incubation (MHI)<br>15 min at 60°C (upper temp. limit), 80% RH | 6 models: 3M 8000, 3M 8210, Moldex 2200, 3M 1860, 3M 1870, and Kimberly Clark PFR95-270 | • For two models (3M 8210 and Moldex 2200), there was a significant reduction in fit; for one model (3M 1860) there was a small increase in odor response. But both effects deemed to be negligible.<br>• 3M 1870 samples experienced a slight separation of the inner foam nose cushion (some to a lesser or greater degree) from the FFR body.<br>• Authors concluded that MHI unlikely to lead to significant changes in fit, odor detection, comfort, or donning difficulty. |
| Lore 2012<sup>53</sup> | Moist heat incubation<br>Uncertain temperature, but likely 65°C for 20 min, unknown RH | 3M 1860s, 3M 1870 | • There was no significant decrease in filter performance |
| Liao 2020<sup>41</sup> | Dry heat<br>20 cycles of 30 minutes at 75°C, unknown RH | 15 x 15 cm pieces of meltblown fabric, described as the most important layer of N95 FFRs<sup>41</sup> | • This is an unpublished report that has not been peer-reviewed, with poorly described methods.<br>• Unchanged filtration efficiency after 15 cycles; very minor decrease after 20 cycles. |
|  | Steam treatment with boiling water vapour (i.e. ~100°C)<br>10 minutes | 15 x 15 cm pieces of meltblown fabric, described as the most important layer of N95 FFRs<sup>41</sup> | • This is an unpublished report that has not been peer-reviewed, with poorly described methods.<br>• No change in filtration efficiency after 3 cycles<br>• Drop in filtration efficiency (from ~97% to ~85%) after 5 cycles, explained by the authors as due to loss of static charge of the fibers. |

FFR, filtering facepiece respirators; MHI, moist heat incubation; RH, relative humidity.

Given this lack of evidence, clinicians have applied a WIWI (would-I-wear-it) test to the process for developing protocol recommendations. Further, based on the literature that was examined during the preparation of this manuscript, the proposed methodology would most likely achieve disinfection against other pathogenic organisms.

As previously mentioned, PPE that are obviously soiled with organic matter should not be reused, as their disinfection is more difficult to achieve using procedures that would not damage them.

Based on the available evidence, the following disinfection steps are proposed, as outlined in Figure 1.

**Figure 1.**
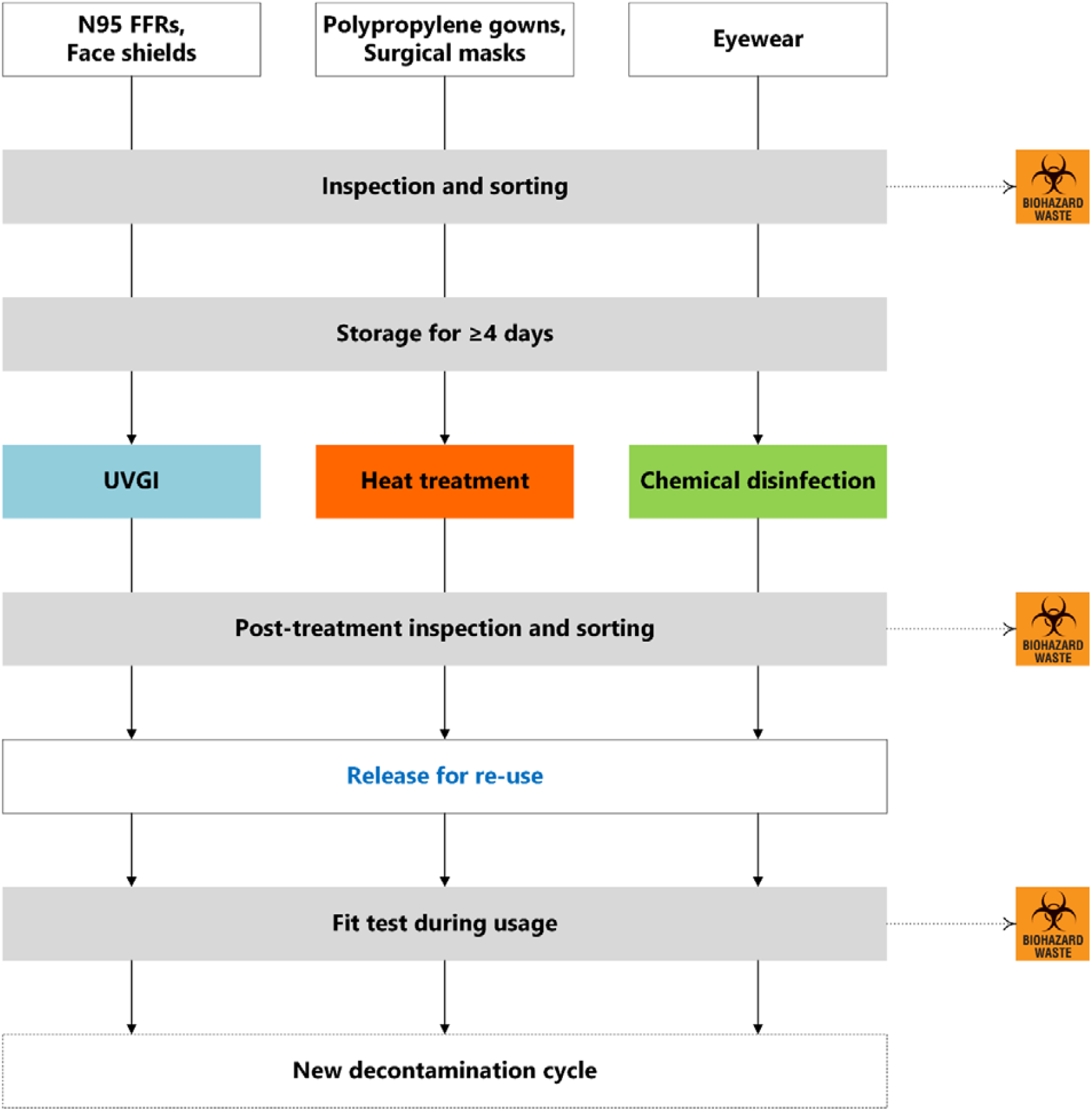
Proposed steps for disinfection from SARS-CoV-2 and re-use of PPE. Dotted lines represent the path (i.e. biohazard waste) for PPE with any sign of damage.

At point of doffing PPE, wearer is to remove and inspect items, looking for any damage or soiling (e.g. blood-stains or presence of organic material). If the PPE is damaged or visibly contaminated, this is to be placed in a bin for biohazard waste. If not damaged or contaminated, PPE is to go into a separate clearly marked bin for re-use. This PPE to be bagged and transported in bin to storage area, where the process outlined below will begin.

### 1. Inspection and sorting

further careful inspection of PPE (including straps); any soiled and damaged PPE to be discarded, intact PPE to be stored.

### 2. Storage

All intact PPE to be stored for at least 4 days in a specially designated area (if conditions can be determined, we suggest ∼20°C and 40–50% relative humidity), ensuring no direct contact between items, and minimizing any creases in material. Locally this will be rooms or enclosures where gowns and other PPE can be hung in four rotating enclosures/rooms, and left for four days.

### 3a. UVGI

After completion of the mandatory stand-down storage period, N95 FFRs and plastic face shields to be treated in the designated UVGI chamber at an applied dose of 2,000 mJ/cm^2^, with this dose applied to each side, i.e. wearer-facing and outer sides.

### 3b. Heat treatment

After completion of the mandatory stand-down storage period, polypropylene gowns and surgical masks to undergo heat treatment at 60°C for 90 minutes, possibly at moderate relative humidity (40–50%).

### 3c. Chemical disinfection

After completion of the mandatory stand-down storage period, eyewear must be disinfected with the appropriate high-grade agent, either through soaking or use of appropriate wipes.

### 4. Re-inspection and sorting

After UVGI, heat treatment, or chemical disinfection, careful re-inspection of PPE (including straps) must take place; any PPE with any sign of damage must be discarded; intact PPE to be packaged for re-use, after being appropriately marked as PPE derived from disinfection, including the number of the disinfection cycle.

### 5. Fit test

Frontline staff to ensure that any decontaminated PPE fit properly as new; at any sign of suboptimal fit, decontaminated gear to be immediately discarded.

Afterwards, a new disinfection cycle to begin.

It should be noted that N95 FFRs should probably be discarded after the fifth re-use. An exception to this rule would be under extreme circumstances, where the alternative to further re-use of suboptimal PPE would be not wearing any protection at all.

## 8. CONCLUSIONS

This protocol provides recommendations for a pragmatic disinfection process for all PPE, that could be rapidly implemented, based on best available evidence. A double-hit process has been proposed due to the immediate urgency of the issue in the current pandemic. Testing of this protocol is in planning stages, but its conservative double-hit approach would most likely achieve disinfection. Based on a total estimated 10% loss of N95 FFRs over 5 cycles, this procedure would increase supply by 400%. We are currently finalizing the tests of the prototype of a UV chamber that would be able to treat a batch of N95 FFRs at the required does in less than 4 minutes.

Careful design of heat chambers and UVC cabinets for re-use of PPE will not only address the problem of short-term supply in the frontline during the pandemic, but also likely lead to considerable cost-savings in the long term. Further, it would also improve the environmental footprint of a given healthcare facility allowing for long-term reuse of PPE, as according to estimates from US hospitals for example, 5.17 tons of waste are generated per staffed bed every year^50^. It is intended that results of protocol testing will be made available as soon as feasible. It is the right of every healthcare worker responding to the current pandemic to have PPE available not only for their protection, but also to reduce the spread of COVID-19^51^.

Acknowledgements

We are grateful to Dr William Lindsley (US CDC) for providing very helpful information during the preparation of this manuscript. Thanks to Dr Lesley Voss (Infectious Diseases, Starship Children’s Health), Dr Emma Best (Department of Paediatrics, University of Auckland), and Professor Cameron Grant (Department of Paediatrics, University of Auckland) for their critical review of initial protocol. Also to Sean McNulty and Kevin Mole (UV Solutionz), Geoff Ray (Engineering Works Supervisor, Taranaki District Health Board), and Anthony Valvoi (Sterile Services Coordinator, Taranaki District Health Board) for their assistance with the site design and UVC testing.

## Data Availability

There are no relevant data in this study that could be made available.

